# Extending Healthcare Access via Telemedicine in Public Libraries: A Mixed Methods Study

**DOI:** 10.1101/2022.08.16.22278852

**Authors:** Pamela B. DeGuzman, Jennifer Garth, Kamya Sanjay, Rebekah M. Compton

## Abstract

**Background:** Despite the potential for telemedicine in public libraries to expand healthcare access to those living a long distance from care and in broadband poor areas, there are few collaborations between libraries and healthcare providers offering these programs.

**Purpose:** To explore licensed independent providers’ perspectives (LIPs) on telemedicine in public libraries as a method of improving equitable access to care for populations lacking the ability to connect to a telemedicine video visit from home.

**Methods:** We used a two-phase explanatory sequential mixed methods design with a quantitative strand followed by a qualitative strand to explore LIPs’ perspective on telemedicine in public libraries. Surveys were analyzed descriptively and to determine group differences. Survey respondents were recruited to participate in interviews, which were analyzed thematically using descriptive content analysis.

**Findings:** Fifty LIPs responded to the survey, and 12 were interviewed. Respondents were overwhelmingly supportive of telemedicine in public libraires, describing how video visits could help multiple vulnerable populations connect to providers when travel was cost prohibitive. They emphasized how connecting at-risk populations to a video visit instead of a telephone call allowed for a more thorough and accurate assessment. While several LIPs were concerned with privacy, others considered a library to be more private than the home. Interviews revealed how chronic illness management may be the ideal visit type for public library-based telemedicine.

**Conclusions:** Given the importance of expanding access sites for telemedicine, providers should consider partnering with libraries in their catchment areas where broadband access is sparse, and patients must travel long distances to care. Managing chronic illnesses using telemedicine in public libraries may be an important approach toward reducing health disparities in populations who live long distances from care and do not have home-based internet access.

## Introduction

Limitations in broadband connectivity in the U.S. have stifled the ability for residents of many communities to connect to their healthcare providers over a telemedicine video visit (VV).^1^ For example, when providers switched to telemedicine visits during the coronavirus pandemic of 2019 (COVID-19) patients without broadband internet access were unable to participate in a VV from home.^2^ Although this inequity is most apparent in rural areas, urban residents can also have difficulty connecting to telemedicine.^3^ Lack of computer equipment and digital literacy (which typically accompanies poor digital access) further limit telemedicine use.^4–7^ As such, lack of broadband access exacerbates an already inequitable situation in which those who experience difficulty traveling to receive care (i.e., rural populations, those lacking transportation), cannot participate in a VV.^8,9^ Until broadband is broadly and affordably available across the U.S., solutions are needed to ensure equitable access to care.

During COVID-19, a promising solution emerged: A few public libraries began offering spaces for residents to connect to a telemedicine VV.^10–13^ The benefits of public libraries providing space for VVs are multi-fold. Not only can community members access high speed internet with assistance from technologically savvy librarians, but also many small and remote libraries never closed or opened quickly after the initial lockdown, demonstrating how libraries can be instrumental in keeping people connected during a public health crisis.^14^ In rural and remote areas, the travel time to a library can be far shorter than to a provider, particularly for specialty care.^15^ As such, public libraries are emerging as an important link in supporting equitable health access.

Although most librarians favor the idea of telemedicine in public libraries (TIPL), few have adopted the programming.^1^ Recent research suggests that a hallmark of successful TIPL programs is strong library-provider partnerships. Provider support may be critical to adoption, as patients are often directed to these programs through a participating licensed independent provider (LIP) rather than through traditional community-based library marketing channels.^3^ However, to date, there has been no research evaluating how LIPs view TIPL, and if they recognize how these programs help reduce disparities. Thus, the purpose of this research was to explore LIPs’ perspectives on TIPL to improve equitable access to care for underserved populations.

## Methods

We used a two-phase explanatory sequential mixed methods design with a quantitative strand followed by a qualitative strand. This design may be used when researchers wish to explore why quantitative results occurred when no explanatory theory or framework is available.^16^ Moreover when conducting implementation research to evaluate barriers to adoption of health promoting technologies and services, Glasgow and colleagues suggest conducting interviews and distributing surveys to non-participants of the technology.^17^ Accordingly, we first collected quantitative data using closed-ended survey questions, and followed this with qualitative interviews intended to provide a deeper explanation of survey responses. The research was approved by the University of Virginia Institutional Review Board for Social and Behavioral Research. All data were collected between May and August 2021.

### Quantitative Methods

We recruited LIPs including physicians, nurse practitioners (NPs), physician assistants, nurse midwives and clinical nurse specialists through multiple channels to gain broad participation. We emailed the survey to four statewide LIP groups and sent survey links via social media through statewide nursing and medical agencies, and encouraged sharing of the survey through snowball sampling. Of note, although we primarily recruited through healthcare provider agencies and associations in one U.S. state, any LIP from any practice setting or geographical location was eligible to participate.

The quantitative survey included information about the research study and 8 questions. The first four were about the provider’s practice (health provider role, practice environment, care delivery model, and patient population), and the last four were about their perspectives on TIPL, specifically support for and concerns with such programs, what types of services they envisioned TIPL being appropriate for, and any perceived barriers to TIPL. A copy of the survey and permission to use it is available by request from the corresponding author. All data were analyzed descriptively. Inferential analysis using chi-square was conducted to determine differences between survey responses by categories with sufficient group sizes.

### Qualitative Methods

Eligibility for the qualitative strand was the same, and participants were recruited through the quantitative survey and through snowball sampling. At the end of the survey, participants were asked to provide follow up information if they were interested in participating in an in-depth interview, and at the end of each interview, participants were asked to share information about the study with colleagues. All participants who provided follow-up contact information or who contacted the principal investigator via email were scheduled for an interview.

The interview guide (**Figure 1**) was designed to illuminate findings of the quantitative strand with more precise data. First, we asked questions about the provider’s practice area and community served. Next, we investigated items revealed through the survey as important to respondents, specifically to uncover drivers of support for TIPL and to gain a clearer understanding of concerns and perceived barriers.

**Figure 1:**
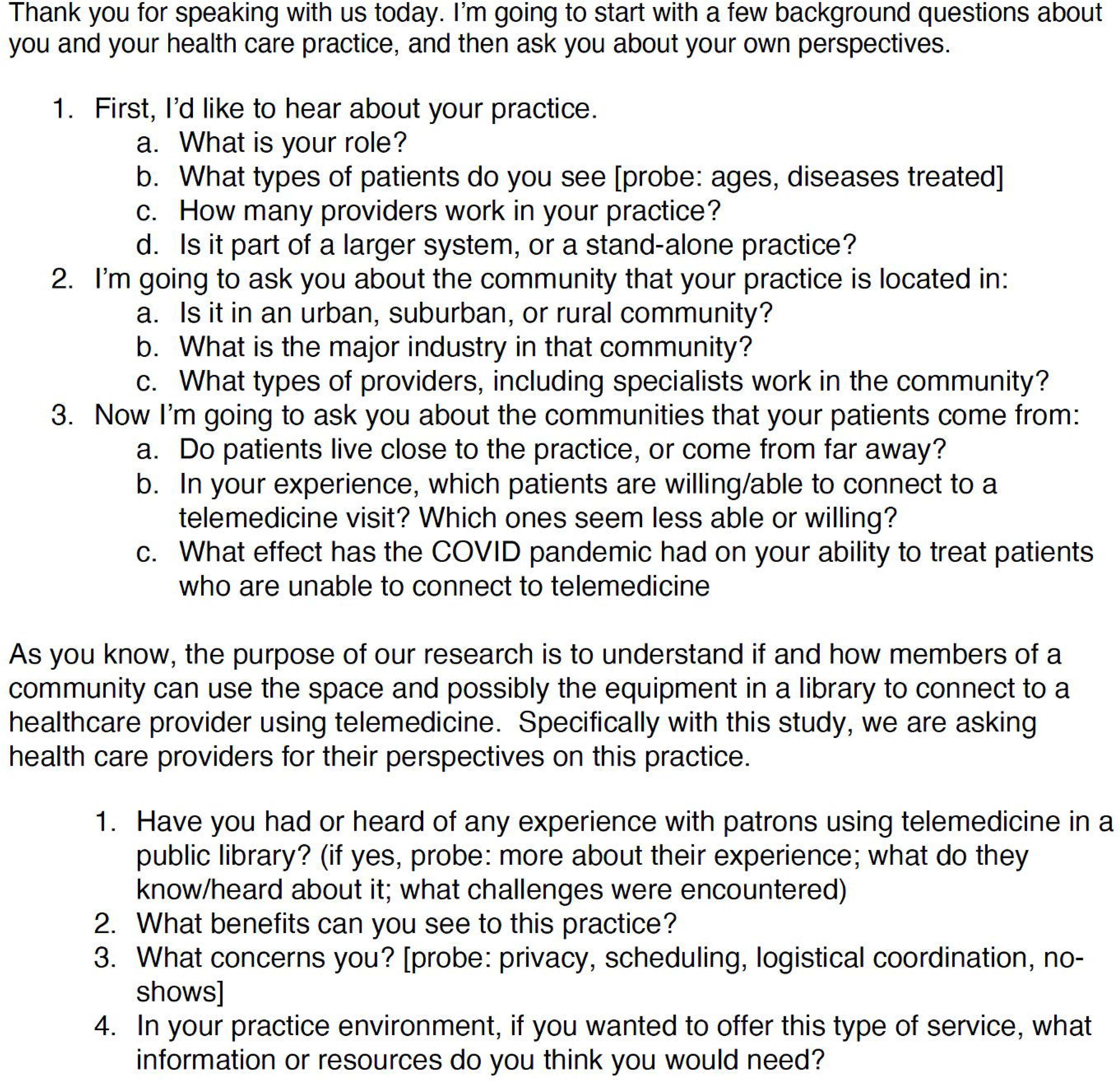
Semi-structured interview guide

All interviews were recorded using Zoom videoconferencing (Zoom Videoconferencing Inc.; v.5.10.4). Verbal consent was obtained prior to recording. Interviews were transcribed verbatim. Once transcriptions were checked for accuracy, the original interviews were deleted. Identifying information was removed and transcriptions were uploaded into Dedoose (v.9.0.17, Los Angeles, CA: SocioCultural Research Consultants, LLC).

We used an inductive, descriptive, qualitative approach to analyze data and reach saturation,^18^ as guided by the study aim. One researcher (J.G.) read through the entire data set multiple times to familiarize themselves with the data prior to coding, then coded all data. A second researcher (P.D.) reviewed the codes and the two discussed and resolved coding discrepancies collaboratively. Codes were collapsed into broader categories, and related categories into themes, which were validated by three members of the research team (P.D., J.G., and K.S). After reaching data saturation, we used the final three interviews to verify findings.^19^

## Results

### Quantitative results

Fifty providers completed the survey. **Table 1** contains a full description of participants’ practice characteristics. The majority of participants were NPs (36/50; 72%), practiced in an outpatient environment (46/50; 92%), worked in primary care (36/51; 72%) and cared for adult patients (29/50; 58%).

**Table 1:**
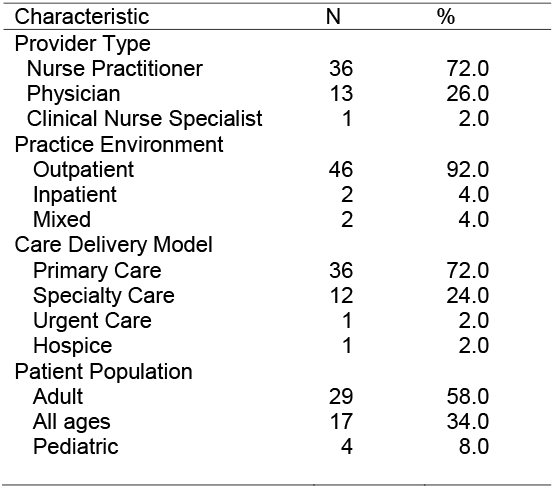
Participant Practice Characteristics (n=50)

**Table 2** contains responses to the survey questions for all respondents, categorized by provider type. Eighty-two percent of providers reported being supportive of TIPL. The largest concerns with TIPL were privacy and security (68%) and patient familiarity with technology (44%). Providers were most favorable of conducting health education using TIPL (52%), followed by health promotion/disease prevention (48%) and chronic illness management (48%). Few providers identified barriers to TIPL; the most common was lack of internet access (20%) followed by inaccurate diagnoses (16%).

**Table 2:** Perspectives on TIPL Organized by Provider Type (n=50)

|  | All (n=50) | Physician<br>(n=13) | Nurse<br>Practitioner<br>(n=36) | Clinical Nurse<br>Specialist<br>(n=1) | p-value |
| --- | --- | --- | --- | --- | --- |
| Open to TIPL |  |  |  |  |  |
| Yes | 42 (82.4) | 12 (92.3) | 29 (80.6) | 1 (100.0) | 0.828 |
| Maybe | 1 (2.0) | 0 (0.0) | 1 (2.8) | 0 (0.00) |  |
| No | 7 (13.7) | 1 (7.7) | 6 (16.7) | 0 (0.00) |  |
| Concerns |  |  |  |  |  |
| Privacy or security | 34 (68.0) | 12 (92.3) | 21 (58.3) | 1 (100.0) | 0.025* |
| Patient familiarity with technology | 22 (44.0) | 10 (76.9) | 12 (33.3) | 0 (0.0) | 0.007* |
| Connectivity issues | 15 (30.0) | 7 (53.8) | 7 (19.4) | 0 (0.0) | 0.019* |
| Malpractice or liability | 13 (26.0) | 5 (38.5) | 7 (19.4) | 0 (0.0) | 0.172 |
| Reimbursement or billing | 6 (12.0) | 3 (23.1) | 3 (8.3) | 0 (0.0) | 0.165 |
| Other | 2 (4.0) | 1 (7.7) | 1 (2.8) | 0 (0.0) | 0.443 |
| Services |  |  |  |  |  |
| Health Education | 26 (52.0) | 12 (92.3) | 12 (33.3) | 1 (100.0) | <.001* |
| Health Promotion/Disease Prevention | 24 (48.0) | 12 (92.3) | 11 (30.6) | 1 (100.0) | <.001* |
| Chronic illness management | 24 (48.0) | 12 (92.3) | 10 (27.8) | 1 (100.0) | <.001* |
| Health Screening | 20 (40.0) | 11 (84.6) | 7 (19.4) | 1 (100.0) | <.001* |
| Other | 4 (8.0) | 2 (15.4) | 2 (5.6) | 0 (0.0) | 0.267 |
| Barriers |  |  |  |  |  |
| Lack of internet access | 10 (20.0) | 4 (30.8) | 6 (16.7) | 0 (0.0) | 0.280 |
| Inaccurate diagnoses | 8 (16.0) | 4 (30.8) | 4 (11.1) | 0 (0.0) | 0.100 |
| Relationship with provider | 4 (8.0) | 2 (15.4) | 2 (5.6) | 0 (0.0) | 0.812 |
| Other | 4 (8.0) | 2 (15.4) | 2 (5.6) | 0 (0.0) | 0.267 |

Note: TIPL = Telemedicine in Public Libraries. Other = Urgent Care and Hospice. P-value is calculated for the difference between nurse practitioners and physicians using Chi-Square analysis. Other services include MAT therapy for low-risk patients, mental health counseling, (n=2), acute care for appropriate issues (n=1). Other barriers include transportation to the library (n=3), lack of devices to use (1), privacy in the library (n=1), lack of ability to use the technology without assistance (n=1). Other concerns include limited broadband in rural areas (n=1), ease of traveling to a library compared with traveling to the physician's office (n=1).
\* p<.05

Because the majority (98%) of providers were either physicians or NPs, inferential analysis was conducted to determine differences between these wo groups. Physicians had greater concerns about privacy and security in the library (92.3% vs 58.3%, p=0.025), patient familiarity with technology (76.9% vs 33.3%, p=0.007), and connectivity issues in the library (53.8% vs 19.4%, p=0.019). Physicians were more supportive of providing telemedicine in the library than NPs for health education (92.3% vs 33.3%, p<.001), health promotion/disease prevention (92.3% vs 30.6%, p<.001), chronic illness management (92.3% vs 27.8%, p<.001) and health screening (84.6% vs 19.4%, p<.001). There were no differences in the barriers identified to TIPL.

**Table 3** contains responses to the survey questions categorized by care delivery model (i.e., primary care, specialty care, hospice, and urgent care). Due to low numbers of hospice and urgent care providers (n=1 each, respectively), inferential statistics were calculated between primary and specialty care. There were no significant differences between groups.

**Table 3:** Perspectives on TIPL Organized by Care Delivery Model (n=50)

|  | Primary<br>Care<br>(n=36) | Specialty<br>Care (n=12) | Other<br>Model<br>(n=2) | P-value |
| --- | --- | --- | --- | --- |
| Open to TIPL |  |  |  |  |
| Yes | 32 (88.9) | 9 (75.0) | 1 (50.0) | 0.393 |
| Maybe | 0 (0.0) | 1 (8.3) | 0 (0.0) |  |
| No | 4 (11.1) | 2 (16.6) | 1 (50.0) |  |
| Concerns |  |  |  |  |
| Privacy or security | 24 (66.7) | 8 (66.7) | 2 (100.0) | 1.000 |
| Patient familiarity with technology | 18 (50.0) | 4 (33.3) | 0 (0.0) | 0.316 |
| Connectivity issues | 10 (27.8) | 3 (25.0) | 1 (50.0) | 0.851 |
| Malpractice or liability | 9 (25.0) | 3 (25.0) | 0 (0.0) | 1.000 |
| Reimbursement or billing | 4 (11.1) | 2 (16.7) | 0 (0.0) | 0.614 |
| Other | 2 (5.6) | 0 (0.0) | 0 (0.0) | 0.404 |
| Services Amendable to TIPL |  |  |  |  |
| Health Education | 20 (55.6) | 5 (41.7) | 0 (0.0) | 0.404 |
| Health Promotion/Disease Prevention | 19 (52.8) | 5 (41.7) | 0 (0.0) | 0.505 |
| Chronic illness management | 16 (44.4) | 3 (25.0) | 0 (0.0) | 0.233 |
| Health Screening | 19 (52.8) | 4 (33.3) | 0 (0.0) | 0.243 |
| Other | 4 (11.1) | 0 (0.0) | 0 (0.0) | 0.228 |
| Perceived Barriers |  |  |  |  |
| Lack of internet access | 6 (16.7) | 3 (25.0) | 1 (50.0) | 0.522 |
| Inaccurate diagnoses | 5 (13.9) | 3 (25.0) | 0 (0.0) | 0.371 |
| Relationship with provider | 4 (11.1) | 0 (0.0) | 0 (0.0) | 0.228 |
| Other | 4 (11.1) | 0 (0.0) | 0 (0.0) | 0.228 |

Note: TIPL = Telemedicine in Public Libraries. Other Model= Urgent Care and Hospice. P-value is calculated for the difference between nurse practitioners and physicians using Chi-Square analysis. Other services include MAT therapy for low risk patients, mental health counseling, (n=2), acute care for appropriate issues (n=1). Other barriers include transportation to the library (n=3), lack of devices to use (1), privacy in the library (n=1), lack of ability to use the technology without assistance (n=1). Other concerns include limited broadband in rural areas (n=1), ease of traveling to a library compared with traveling to the physician's office (n=1).

### Qualitative results

Twelve providers agreed to an interview. All 12 were NPs. To preserve anonymity and aid analysis, NPs are identified in the results as working in either primary (n=7) specialty (n=4), or psychiatric care (n=1). Three worked in a rural setting, five in a suburban setting and four in an urban setting. Two worked primarily with pediatric patients, 5 with all ages and 5 primarily with adults. Four themes emerged from the qualitative data: *improving access for multiple at-risk populations, privacy concerns, chronic illness management as an ideal public library telemedicine visit*, and *providers’ reliance on visual aspects of telemedicine*.

#### Theme 1: Improving Access for Multiple At-Risk Populations

Providers identified populations that could benefit from the improved access that TIPL setting offers, including those with long travel distances, transportation barriers, limited internet access, and limited digital skills; the uninsured and underinsured; and caregivers of young children. Rural patients and the underinsured were among those often noted as having long travel distances and limited transportation options. A primary care NP working in a suburban community commented on the long driving time for their rural patients: “Round trip is probably four hours.” A rural primary care NP who often referred their clientele to a regional academic center for specialty care noted the travel difficulties their patients experienced, leading to missed appointments: “[Planning for the trip begins] five days ahead to get set up with transport …[then] you’re looking at the transport not showing up missing the specialty consultation that they’ve been waiting months for.” NPs serving a suburban and urban clientele reported fewer issues with transportation to appointments. One primary care NP stated, “Most of our patients have…independent, easy access to their own personal transportation.” However, transportation difficulties were not exclusive to rural areas. One primary care NP working in a suburban area who treated a predominantly uninsured and underinsured population described most patients as having to take “two and three buses” to attend their appointments.

NPs serving rural populations recognized how libraries could help their clientele lacking home-based broadband internet connect to a telemedicine. Several primary care NP described the difficulties rural patients had connecting to the internet for telemedicine visits. According to one, “If you do not live in a neighborhood with Comcast or FiOS, you only have satellite…the connection is very poor. And then if people are dependent upon their phones, for mobile access to internet, the connections did not [always] work.” Another primary care NP reported the same experience with patients attempting to connect: “We have had some access issues of patients that are in rural, or more remote locations…actually connecting to the internet [but then] having connection issues.” A specialty care NP with a surgical practice envisioned how their rural patients could utilize TIPL for post-operative visits, stating: “if they don’t have Internet at their rural house…[they] can drive 10 minutes to the library or five minutes and still get that that video call in.”

An NP working in an urban setting stated that libraries may be useful for patients who “are unsure of [how to use] their phone or their computer” because libraries can help them understand how to use the technology. A primary care NP commented on how the library could help bridge the gap to support telemedicine for the digitally underserved: “[The] library is genius because of the internet connection.” Similarly, a specialty care NP stated: “being able to use, like, newer technology at the library, equipment, internet is I think the big [benefit].”

NPs identified how caregivers of young children could benefit from telemedicine visits in the library. One primary care NP serving a rural population stated, “a lot of [patients] want to continue seeing us over telehealth…because they still have kids [at home].” A specialty care NP identified that parents and caregivers lacking supervision for their children could benefit from a place where children have access to activities while the parent is engaged in the visit: “I’ve been [the provider] on visits before where there’s five kids bouncing around in the background, and I’m trying to teach things [to the parent or caregiver]…And so you might be better even just to be able to put your other two kids in the reading circle or whatever, right?”

#### Theme 2: Privacy Concerns

NPs across multiple practice types and settings expressed concerns about the ability for libraries to provide adequate privacy for a health visit. One primary care NP serving a rural population stated that they would want “assurance that patients know that nobody in the library can hear what they’re saying.” They further described how patients might also have similar concerns. “You might see some hesitation unless there is an established channel…It would need to be a space that for HIPAA regulations could be pretty confined.” A psychiatric NP working in addictions medicine in a suburban community described similar, but specialty-specific concerns: “There’s another layer of privacy on top of psychiatry, which is super private anyway. …The ability to openly communicate is concerning. It’s already concerning [depending on] who’s in the background of their home…Add strangers [in the library] …that’s very concerning to me.” A specialty care NP working in an urban setting doubted that their patients would trust the library as a healthcare setting: “Some of my patients are very private…would have nothing, want nothing to do with a public library.”

Despite these concerns, two NPs viewed the library as a place where privacy could be *enhanced* compared with a home visit. A specialty care NP working in an urban setting stated, “A public library would actually probably be more secure than what I’ve seen. Even within the home, there are people coming and going and interruptions and things.” They further described how libraries accommodate privacy: “They have these little rooms that you can reserve… Something like that would be an awesome thing to offer.” Another NP with knowledge of library spaces commented on the ability for patrons to utilize a private room stated, “A lot of libraries have community rooms, you know, and that’s the ideal place to do some of this.”

#### Theme 3: Chronic Illness Management as Ideal Public Library Telemedicine Visits

Most visits providers described as appropriate for TIPL involved chronic illness management. Providers described the usefulness of telemedicine for management and education visits, if visits did not require physical palpation. A pediatric primary care NP described how as their office began to open up for more in person visits after the initial shut down of COVID-19, they planned to keep many follow-up appointments online for particular groups of patients including, “every three to six months check ins [for patients taking] anxiety medication or depression, or [those] managing behavioral concerns, ADHD…We can follow up with them more frequently and regularly and easier [using telemedicine].” Another primary care NP provider commented on the use of telemedicine for their psychiatric visits: “I find that televisits [sic] lend well to psych follow ups… ‘everything’s fine I just need my medication refills.’ This is just a routine follow up.” Another pediatric primary care NP described the usefulness of telemedicine for managing certain illnesses, including “asthma management, depression management… a lot of those things you can do over a televisit that the population really needs and you know, they don’t ever come back until they need that next refill in three months or a year.”

NPs discussed how libraries could be enhance a predominantly education-based visit. A specialty care NP noted the benefit of conducting an education visit in the library prior to a scheduled procedure because of the potential to disseminate educational material electronically to patients. “If I could fax something, or email something to be printed off at the library, that might be good, because [I give patients] a lot of educational material.” Further, a primary care NP serving a rural population described how patients could take advantage of libraries’ place as source of educational information for community members lacking internet access: “It’s a place where patients are getting resources already. They’re often going and looking up things if they don’t have a computer.”

NPs noted some visits that would not be conducive to either telemedicine or the public library setting, either due to limited assessment when relying on remote care or the need for high levels of privacy. A primary care NP stated, “acute visits are absolutely terrible. Anything…respiratory-related or skin-related [can lead] to deviation from the standard of care.” A specialty care NP serving a pediatric population explained, “There are some exam portions that you might do at home that you might not do in a library.”

#### Theme 4: Providers’ Reliance on Visual Aspects of Telemedicine

Providers in our study discussed the benefits of the visual aspect of a telemedicine visit compared to a phone call for assessment. A specialty care NP commented that being able to “see the person is invaluable.” One provider stated a preference for video rather than relying on the patient’s description: “I always find that even talking on the phone, you kind of get half of the picture. But then when I can get that video, I can see the patient, I can see that they’re not in distress, they’re sitting there, they don’t look like they’re wincing in pain.” Likewise, a primary care NP provider serving a rural community described how a VV allowed them to assess the environment, stating “[I can] actually see what’s going on in their homes. I can see how they look in their own environment, … with their overall being, a little bit better than when they take a shower and come and see me in the office and look all so well.”

Additional benefits of VV identified by NPs were the enhanced ability to develop a therapeutic relationship and diagnose patients appropriately. A psychiatric NP described the ability to see the patient’s facial expressions as a critical visit component. “[When] you’re working in psychiatry…in that small setting of just needing to talk to somebody in the room and you can’t see facial expressions. That’s hard.” A primary care NP stated a preference for video visits because they felt it increased the ability to ensure an accurate diagnosis, noting, “patients do a lot of self-diagnoses with telephone calls.”

## Discussion

### Support for Telemedicine in Public Libraries

Over 80% of LIPs surveyed supported the idea of patients connecting to a telemedicine video visit from a public library. Interviews indicated that support was driven chiefly by their experience with patients who regularly encountered barriers both connecting to a VV from home and travelling to appointments, both well-known barriers for those with lower incomes, the underinsured and uninsured.^5,15,20^ Of note, in the U.S., those lacking home-based broadband internet are more likely to be poor, have lower education, are less likely to have health insurance, and more likely to be disabled and have a shorter life expectancy.^21^ While little is yet known about patients who use TIPL, there is much evidence to suggest that libraries target health-related programming toward similarly vulnerable populations.^22^ As such, expanding TIPL programs represents an important step toward equalizing access for those already struggling to maintain health.

Providers also suggested they supported TIPL because of the benefit of visualizing the patient, compared to an audio-only visit. However, without alternative access points, many patients will be left with telephone as the only remote option, which our study suggest leads to a poorer quality visit, thus broadening the digital health divide. In the U.S. computer ownership is lower among rural residents, older persons, disabled persons, and those with lower incomes and education.^7^ While one can connect to a visit from a smartphone, 20% of those living in the rural U.S. do not own one, and only 72% of rural residents have home broadband. ^23^ In many rural communities, libraries are the only accessible place where residents can connect to broadband internet.^24^ Furthermore, rural residents are less likely to have digital skills and libraries offer the added benefit of technology-savvy librarians who can assist with navigating digital equipment platforms needed for telemedicine.^25^

### Addressable Concerns

Both qualitative and quantitative data indicated that ensuring privacy during a TIPL visit was the top concern among providers, followed by patients’ ability to use technology to connect to a visit. Providers may lack familiarity with the privacy and technology support available for telemedicine at modern public libraries which include standalone soundproof kiosks, mobile libraries, white noise machines, and exclusive use of private meeting rooms.^3^ Finally, it is important to note that not all NPs viewed privacy in the library as a concern. Two NPs stated that the library would be *more* private than patients’ homes. The potential for enhanced privacy is particularly relevant in situations where overheard communication may place a patient at risk from someone in their own home (such as with someone who is subject to violence at home). In this case, using the internet to connect to a VV with provider *outside* of the home, when unable to attend an in-person visit may increase both access and safety. Providers may be similarly unaware of other services offered at modern libraries that can enhance health and safety, such as social workers who are deployed throughout many public libraries.^26^ A potential solution is for provider practice groups and health systems caring for underserved populations to consider integrating information from libraries in their service area when conducting community health assessments, and involving them as collaborative stakeholders in planning, so that library health programming can be disseminated among all relevant providers.

### Implications for Healthcare Delivery, Research and Policy

Providers who currently offer chronic illnesses management visits over telemedicine may find TIPL to be an ideal way to increase visit attendance for those patients who have difficulty both attending in-person appointments and streaming a telemedicine visit. In the U.S., people who live in communities with lower internet access have significantly higher rates of mortality from cardiovascular disease, cancer, and diabetes.^21^ Offering TIPL as an alternative access point to this population has the potential to improve health and quality of life for the digitally underserved. No research has yet evaluated the impact of TIPL on patient health outcomes, but this is an important direction for future study.^1^ As programs continue to emerge, research is needed that will measure the impact on missed appointments, distance traveled, as well as population-specific health outcomes such as hemoglobin A1C for diabetics or adherence to cancer prevention behaviors for cancer survivors.

This is the first study to document reasons why a video visit may be superior to telephone calls for patient assessment, establishment of therapeutic communication, and diagnosis. This is an important finding in the current policy climate, due to the continued preponderance of telemedicine visits and the current debate over the breadth of reimbursement funding for telemedicine, as insurance companies may soon cease reimbursing non-mental health providers for telephone visits, making improving access to VV that much more critical.^27^ Regardless of funding current changes, research evaluating differences in quality of audio-only and audio-visual visits compared with in-person visits can help support efforts to identify opportunities to reduce access disparities going forward.

## Limitations

This study was conducted with a limited recruitment and a small sample, and may not represent all health provider perspectives. Our survey was answered predominantly by NPs and physicians, and only NPs participated in the interviews, limiting generalizability across LIPs. Only providers from a few specialty areas were represented in the interviews and given the potential for TIPL to impact specialized chronic illness management, future studies need to more robustly capture these providers. Because we did not gather data about the providers’ geographic location (other than rurality), and our recruitment was primarily conducted through statewide groups, or sample may be further limited to one geographic are of the U.S. Overall, a larger, more representative study is needed to better identify support for, and concerns with telemedicine programs in public libraries. Finally, although telemedicine familiarity has increased since COVID-19, participants may still \have limited experience with its use.

## Conclusions

Providing support for patients without broadband to connect to a telemedicine VV from alternative locations is critical to reducing health disparities related to the digital divide. Few providers seem to be aware of how these programs can improve access to a technologically supported, private healthcare visit in a public library. TIPL visits may be particularly appropriate for visits aimed at managing chronic diseases. Future research is needed to evaluate how TIPL impacts healthcare outcomes in specific patient populations with limited digital and healthcare access.

## Data Availability

All data produced in the present study are available upon reasonable request to the authors

